# Early-Life Sugar Restriction During the First 1000 Days and Adult Kidney Disease: A Natural Experiment with Phenotypic and Metabolomic Mediation

**DOI:** 10.64898/2026.05.06.26352507

**Authors:** Xianglian Cai, Xiaolong Liang, Dan Chen, Yiwei Zhang, Ziliang Ye, Yanjun Zhang, Sisi Yang, Xiaoqin Gan, Yu Huang, Yiting Wu, Yuanyuan Zhang, Xianhui Qin

## Abstract

**Background:** The first 1000 days from conception to age 2 years represent a critical window for kidney development, during which nutritional exposures may have lifelong programming effects. Whether early-life sugar restriction reduces long-term kidney disease risk remains unknown.

**Methods:** Using the UK sugar rationing policy (1942–1953) as a natural experiment, we compared risks of chronic kidney disease (CKD) and acute kidney injury (AKI) among 64,942 UK Biobank participants born around the rationing period. Duration of early-life exposure was categorised. Cox proportional hazards models estimated hazard ratios (HRs). Negative control analyses included non-UK-born UK Biobank participants and the Chinese CHARLS cohort. Mediation analyses integrated clinical phenotypes and metabolomic profiles.

**Findings:** Compared with never-exposed individuals, those exposed to sugar rationing throughout the first 1000 days (in utero to age 2 years) had lower risks of CKD (adjusted HR 0.78, 95% CI 0.66–0.93) and AKI (0.79, 0.69–0.90). Negative control analyses showed null associations. Mediation analyses indicated that metabolic efficiency (basal metabolic rate), body composition (fat-free mass), and lipid metabolism mediated 4–9% of the protective association. A distinct metabolomic signature characterised by higher polyunsaturated fatty acids and lower VLDL subfractions was identified.

**Interpretation:** Sugar restriction during the first 1000 days is associated with lower risks of CKD and AKI in adulthood, partially mediated by favorable metabolic efficiency, body composition, and lipid profiles. These findings identify early-life sugar exposure as a modifiable developmental programming factor for lifelong kidney health and support public health strategies to reduce added sugar intake during pregnancy and early childhood.

**Funding:** National Natural Science Foundation of China, and others

## Introduction

Chronic kidney disease (CKD) affects an estimated 788 million people globally and was the ninth leading cause of death in 2023. Acute kidney injury (AKI) complicates up to one-third of hospitalizations and accelerates CKD progression [1–3]. Identifying modifiable risk factors during critical developmental windows is therefore a public health priority.

The first 1000 days from conception to age two shape lifelong kidney health [4–6]. Nephrogenesis is completed before birth; nephron endowment—a key determinant of adult CKD risk—is directly programmed by the in-utero metabolic environment and cannot be restored later in life [5,7]. Experimental studies show that maternal high-sugar intake activates the renin–angiotensin–aldosterone system and promotes renal lipotoxicity in offspring [10,35,36]. Yet, public health strategies remain focused on adult lifestyle changes, and human evidence linking early-life sugar restriction to lower CKD and AKI risk is virtually absent.

The UK sugar rationing policy (1942–1953) offers a unique natural experiment. Sugar intake was strictly limited during early life to levels matching current WHO recommendations (<15 g/day for young children), followed by a sharp post-rationing rise [11–15]. Leveraging this design, a growing body of quasi-experimental studies has demonstrated that early-life sugar rationing protects against a wide range of chronic conditions in adulthood. These include diabetes and hypertension [16], cardiovascular disease [17], metabolic dysfunction-associated steatotic liver disease [18], asthma and chronic obstructive pulmonary disease [19], heart failure [20], and anxiety [21]. However, whether early-life sugar restriction directly protects against kidney disease—and through what biological pathways—remains unknown.

Here, we used this quasi-experimental setting to examine the association between early-life sugar restriction and incident CKD and AKI in adulthood. We further combined clinical phenotyping with large-scale metabolomic profiling to identify the metabolic signature of early-life sugar restriction and to quantify potential mediating pathways, including body composition, metabolic efficiency, and circulating metabolites.

## Methods

### Study Design and Participants

The UK Biobank enrolled over 500,000 participants aged 37–73 years between 2006 and 2010 across England, Wales, and Scotland [22–24]. Ethical approval was obtained from the North West Multicenter Research Ethics Committee (11/NW/0382), and all participants provided written informed consent. We identified 74,214 individuals born between October 1951 and March 1956. After excluding 6,742 non-UK-born participants, 896 adopted individuals, 1,629 from multiple births, and 5 pregnant women, the final analytical cohort comprised 64,942 participants (**Figure 1A**). Of these, 49,988 with available plasma metabolomics data were included in secondary analyses.

**Figure 1.**
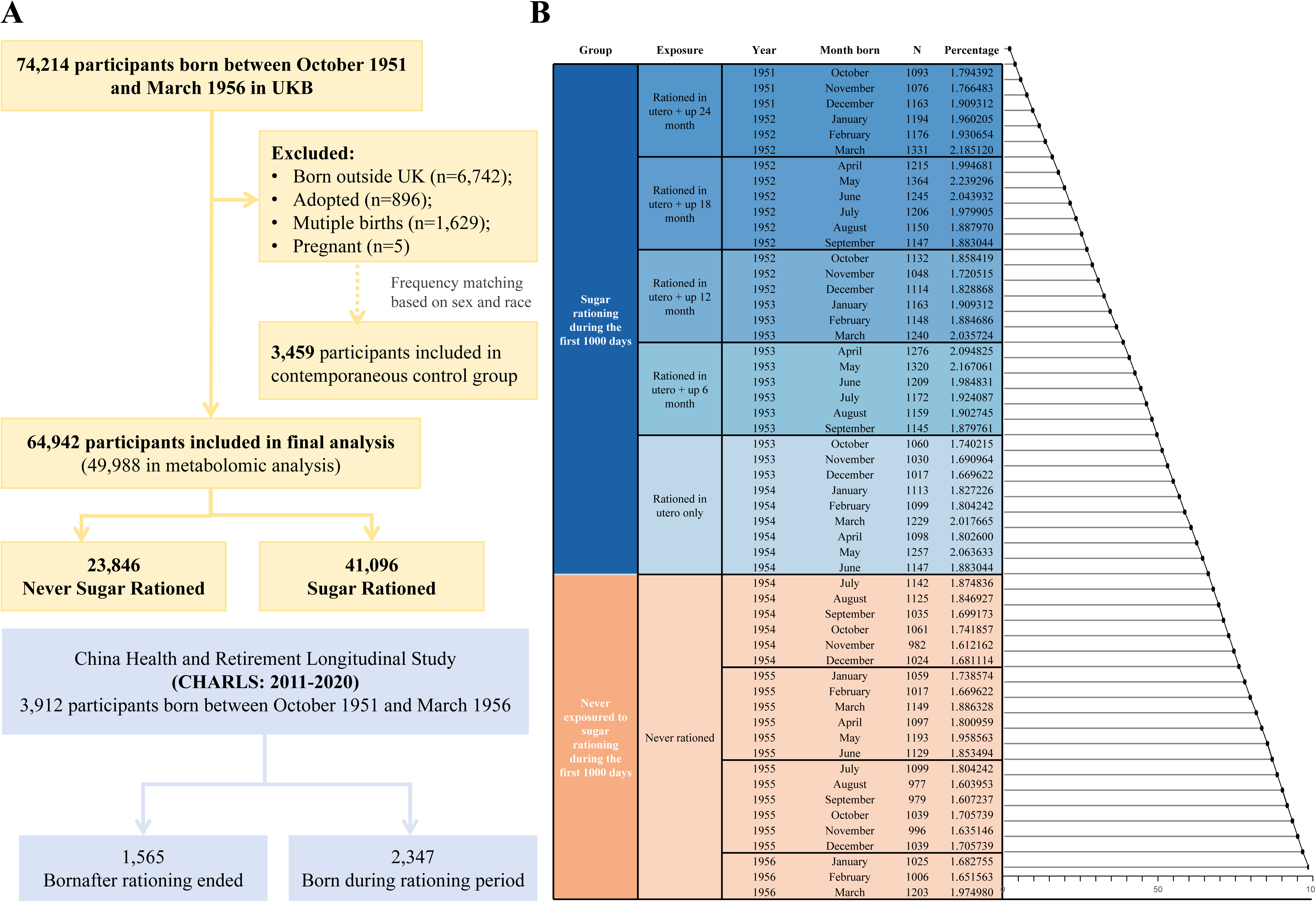
Study Design and Population. **(A)** Flow chart of the study participants. **(B)** Cumulative distribution of births by calendar month and exposure to sugar rationing. Blue represents the sugar-rationed group; orange represents the never-rationed group. The cumulative rate on the x-axis increases from 0% at the top to 100% at the bottom. The first never-rationed group (born July–December 1954) served as the reference in Figure 2. **Abbreviations:** UK, United Kingdom; UKB, UK Biobank.

### Early-Life Exposure to Sugar Rationing

Participants were classified according to their exposure to UK sugar rationing during the first 1000 days of life, defined relative to the cessation of rationing in September 1953. Rationing exposure was categorized as unexposed, exposed in utero only, or exposed in utero plus up to 6, 12, 18, or 24 months postnatally (**Figure 1B**). For main analyses, exposure groups were condensed into four categories consistent with previous reports [16, 17]: unexposed, exposed in utero only, exposed in utero plus the first postnatal year (0–1 years), and exposed in utero plus the first two postnatal years (1–2 years). These cutoffs align with key developmental transitions (weaning and introduction of solid foods) and preserve statistical power [16,17].

### Plasma Metabolite Profiling

Plasma metabolomic data were generated using a targeted high-throughput nuclear magnetic resonance (NMR) platform (Nightingale Health Ltd). The assay quantified 249 metabolic measures (168 absolute concentrations, 81 derived ratios), covering amino acids, fatty acids, lipoprotein subclasses, cholesterol subtypes, and inflammatory markers. All metabolite measures were log-transformed to approximate normality and standardized to z-scores prior to analysis.

### Laboratory Measurements

Blood and urine samples were collected and processed according to standardized protocols [25]. Serum creatinine, urinary albumin, and urinary creatinine were measured at a central laboratory. Estimated glomerular filtration rate (eGFR) was calculated using the CKD-EPI equation [26], and urine albumin-to-creatinine ratio (UACR) was derived from the measured concentrations.

### Covariate Assessment

Baseline characteristics were ascertained through standardized questionnaires. Demographic covariates included age, sex, ethnicity, educational attainment, Townsend Deprivation Index, and parental health. Lifestyle factors included smoking status, alcohol consumption, and physical activity—categorized as optimal if participants engaged in moderate-to-vigorous activity on more than four days per week [27]. Early-life exposures encompassed birthplace, birth season, maternal smoking, and breastfeeding. Geographic birth coordinates were also recorded. History of cardiovascular disease (self-reported or physician-diagnosed stroke, coronary heart disease, or heart failure), hypertension (systolic/diastolic blood pressure ≥140/90 mmHg, self-report, antihypertensive medication, or medical records), and prevalent diabetes (history of diabetes, diabetes medication, or HbA1c ≥6.5%) were defined accordingly. Detailed definitions of phenotypic measures are provided in the **Supplementary Methods**.

Macroeconomic dietary data, including real food prices adjusted for the Consumer Price Index, were sourced from the National Food Survey (1950–1960) [28]. Annual data were previously detailed by Gracner and colleagues [16].

### Polygenic Risk Score Calculation

Polygenic risk scores (PRSs) were computed as follows: a weighted score for higher eGFR (indicating lower genetic risk for CKD) using 263 genome-wide significant SNPs [29], and a score for AKI based on four SNPs [30], with higher scores indicating greater AKI susceptibility. Genotype quality control and imputation have been described previously [31].

### Study Outcomes

The primary outcomes were incident CKD and AKI, ascertained through linkage to hospital inpatient and death registry data. CKD was identified using ICD-9, ICD-10, and OPCS-4 codes [24, 32]; AKI was identified using ICD-10 codes [33]. Detailed code definitions are provided in **Table S1**. Age at onset was calculated as the difference between the date of diagnosis and the participant’s date of birth.

### Contemporaneous Negative Control Analysis

To account for secular trends and unmeasured confounding, we performed negative control analyses in two populations not exposed to UK sugar rationing: non-UK-born UK Biobank participants and participants from the China Health and Retirement Longitudinal Study (CHARLS) [34]. In these unexposed cohorts, birth timing relative to September 1953 does not correspond to variation in early-life sugar exposure; we therefore hypothesized null associations with kidney outcomes.

We identified 3,459 non-UK-born participants frequency-matched (sex and ethnicity) to the UK-born group. For CHARLS, we included 3,912 adults aged ≥45 years from the 2011–2020 waves. In both cohorts, participants were categorized as born before or after the end of sugar rationing (September 1953), mirroring the primary exposure definition. The renal outcome for CHARLS was eGFR <60 mL/min/1.73 m². Follow-up began at birth and ended at the earliest of outcome onset, death, loss to follow-up, or study end. Detailed cohort selection and variable harmonization are provided in the **Supplementary Methods**; participant flow for CHARLS is shown in **Figure 1A**.

### Statistical analysis

Baseline characteristics were summarized as mean (SD) or number (%). Differences between exposure groups (never rationed *vs.* rationed) were assessed using ANOVA or chi-square tests.

Temporal confounding was evaluated in 6-month birth cohorts. Using Cox proportional hazards models, we estimated hazard ratios (HRs) for CKD and AKI by duration of sugar rationing exposure, with participants born July–December 1954 as the unexposed reference. Covariate selection was guided by a directed acyclic graph (**Figure S1**): Model 1 adjusted for sex and ethnicity, and Model 2 additionally adjusted for birth-related factors (birthplace, birth coordinates, birth month), Consumer Price Index (CPI)-adjusted real food prices, survey year, parental history of cardiovascular disease or diabetes, and PRSs for eGFR or AKI. Missing data (**Table S2**) were handled by multiple imputation via the mice package in R.

Unadjusted Nelson–Aalen smoothed hazard curves (**Figure S2**) and Kaplan–Meier survival curves (**Figure S3**) were estimated to describe the unadjusted association between rationing and time to disease diagnosis. Model fit was assessed using Akaike and Bayesian information criteria (**Table S3**); the Gompertz distribution was selected for cross-study comparability [17].

Sensitivity analyses included Gompertz models with Model 2 covariates, additional adjustment for breastfeeding and maternal smoking, further adjustment for adult socioeconomic and lifestyle factors, supplemental adjustment for adult disease status (history of diabetes, hypertension and cardiovascular disease) and competing-risk regression (Fine–Gray) for death. Stratified analyses examined effect modification by sex, ethnicity, birthplace, parental history, and PRS for eGFR or AKI (dichotomized at median).

For mediation analyses, participants with prevalent CKD [29, 35] or AKI [36] at enrollment were excluded. Candidate clinical phenotypes were screened using multivariable linear regression (FDR-corrected *P* < 0.05); only those also associated with kidney disease risk in Cox models were retained. For circulating metabolites, we identified metabolites associated with sugar rationing using LASSO regression with 10-fold cross-validation. Metabolites were grouped into biologically relevant categories; a summary score for each category was derived as a weighted sum using LASSO coefficients. Individual metabolites identified as key drivers were also examined. Mediation analyses were performed using the *mediation* R package with 1,000 bootstrap iterations. All mediation models were adjusted for Model 2 covariates.

For non-UK-born participants and the CHARLS cohort, we fitted both Cox proportional hazards and Gompertz parametric survival models. Non-UK-born analyses retained all primary covariates except birthplace-related variables; CHARLS models were adjusted for sex, nationality, region, birth month, and survey year.

All analyses were conducted using R version 4.1.1. A two-sided *P* < 0.05 was considered statistically significant.

## Results

### Study Population Characteristics

The study cohort included 64,942 participants, of whom 41,096 (63.3%) were exposed to early-life sugar rationing and 23,846 (36.7%) were not. The rationed group was older, more likely to be born in spring, had a higher prevalence of breastfeeding in infancy, and a lower prevalence of parental diabetes (**Table 1**). They also had lower Townsend Deprivation Index scores, lower rates of current smoking, more moderate alcohol consumption, and more history of diseases (**Table S4**).

**Table 1.**
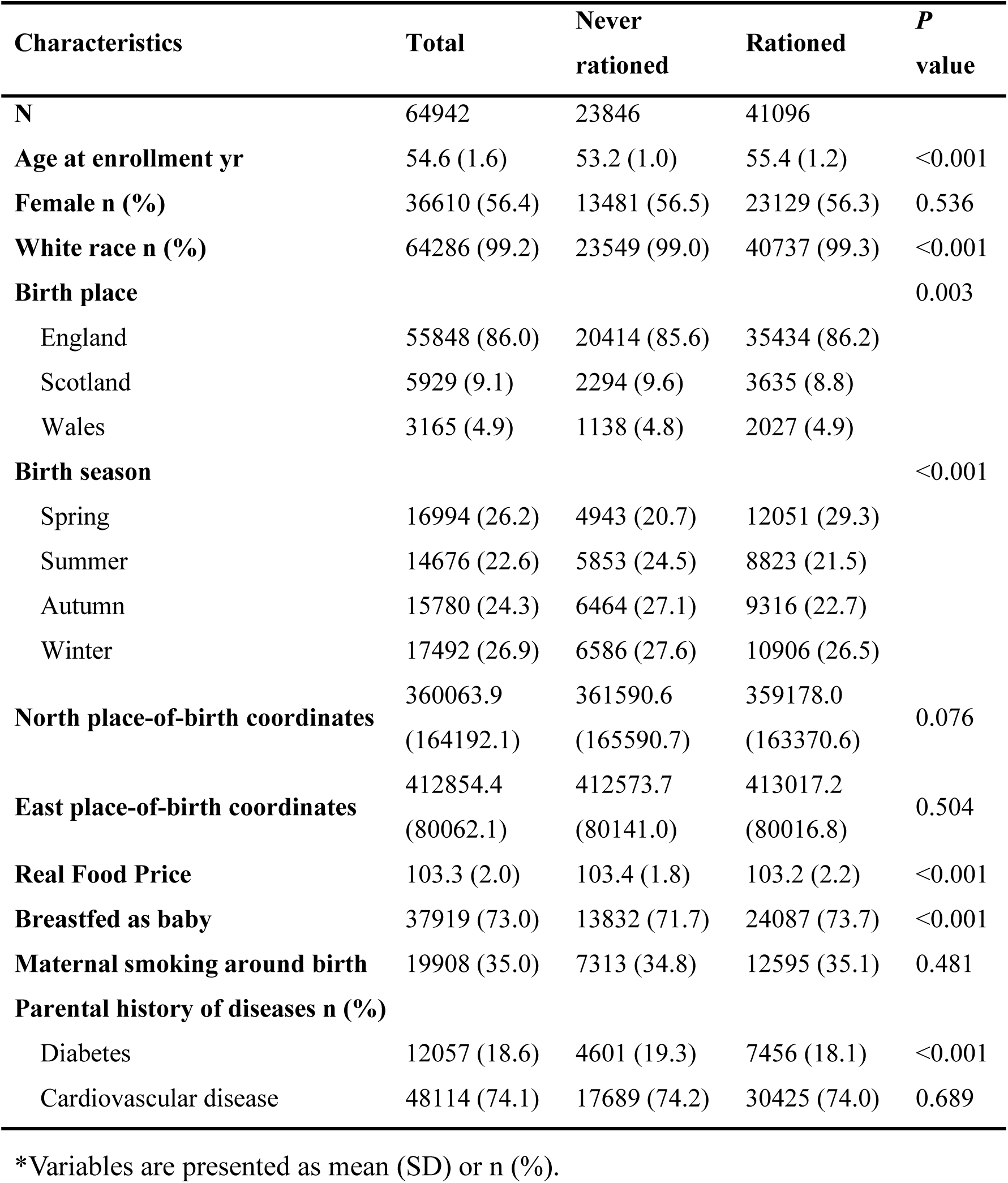
Baseline Characteristics of Study Participants According to Early-Life Exposure to Sugar Rationing*.

| **Characteristics** | **Total** | **Never rationed** | **Rationed** | ***P* value** |
| --- | --- | --- | --- | --- |
| **N** | 64942 | 23846 | 41096 |  |
| **Age at enrollment yr** | 54.6 (1.6) | 53.2 (1.0) | 55.4 (1.2) | <0.001 |
| **Female n (%)** | 36610 (56.4) | 13481 (56.5) | 23129 (56.3) | 0.536 |
| **White race n (%)** | 64286 (99.2) | 23549 (99.0) | 40737 (99.3) | <0.001 |
| **Birth place** |  |  |  | 0.003 |
| England | 55848 (86.0) | 20414 (85.6) | 35434 (86.2) |  |
| Scotland | 5929 (9.1) | 2294 (9.6) | 3635 (8.8) |  |
| Wales | 3165 (4.9) | 1138 (4.8) | 2027 (4.9) |  |
| **Birth season** |  |  |  | <0.001 |
| Spring | 16994 (26.2) | 4943 (20.7) | 12051 (29.3) |  |
| Summer | 14676 (22.6) | 5853 (24.5) | 8823 (21.5) |  |
| Autumn | 15780 (24.3) | 6464 (27.1) | 9316 (22.7) |  |
| Winter | 17492 (26.9) | 6586 (27.6) | 10906 (26.5) |  |
| **North place-of-birth coordinates** | 360063.9<br>(164192.1) | 361590.6<br>(165590.7) | 359178.0<br>(163370.6) | 0.076 |
| **East place-of-birth coordinates** | 412854.4<br>(80062.1) | 412573.7<br>(80141.0) | 413017.2<br>(80016.8) | 0.504 |
| **Real Food Price** | 103.3 (2.0) | 103.4 (1.8) | 103.2 (2.2) | <0.001 |
| **Breastfed as baby** | 37919 (73.0) | 13832 (71.7) | 24087 (73.7) | <0.001 |
| **Maternal smoking around birth** | 19908 (35.0) | 7313 (34.8) | 12595 (35.1) | 0.481 |
| **Parental history of diseases n (%)** |  |  |  |  |
| Diabetes | 12057 (18.6) | 4601 (19.3) | 7456 (18.1) | <0.001 |
| Cardiovascular disease | 48114 (74.1) | 17689 (74.2) | 30425 (74.0) | 0.689 |

\*Variables are presented as mean (SD) or n (%).

### Association between Early-Life Sugar Rationing and Risk of Kidney Diseases

During follow-up, 1,055 participants developed CKD and 1,688 developed AKI. Disease risk increased with age in both groups, but more steeply among the never-rationed group. Divergence in risk began around age 50, with the largest differences observed after age 60 (**Figure S2**). Unadjusted Kaplan-Meier survival curves showed progressively lower cumulative incidence with longer rationing exposure (**Figure S3**).

Using participants born July–December 1954 as the unexposed reference, those exposed in utero and beyond 12 months had a 29% lower risk of CKD (adjusted HR = 0.71, 95% CI: 0.50–1.00; *P* =0.049; **Figure 2A, Table S5**). Longer postnatal exposure (>18 and >24 months) similarly conferred lower CKD risk. For AKI, a comparable protective trend was observed, with statistical significance for in utero exposure exceeding 12 months (HR = 0.75, 95% CI: 0.57–1.00; **Figure 2A, Table S5**). Results from Gompertz models were consistent (**Figure S4**).

**Figure 2.**
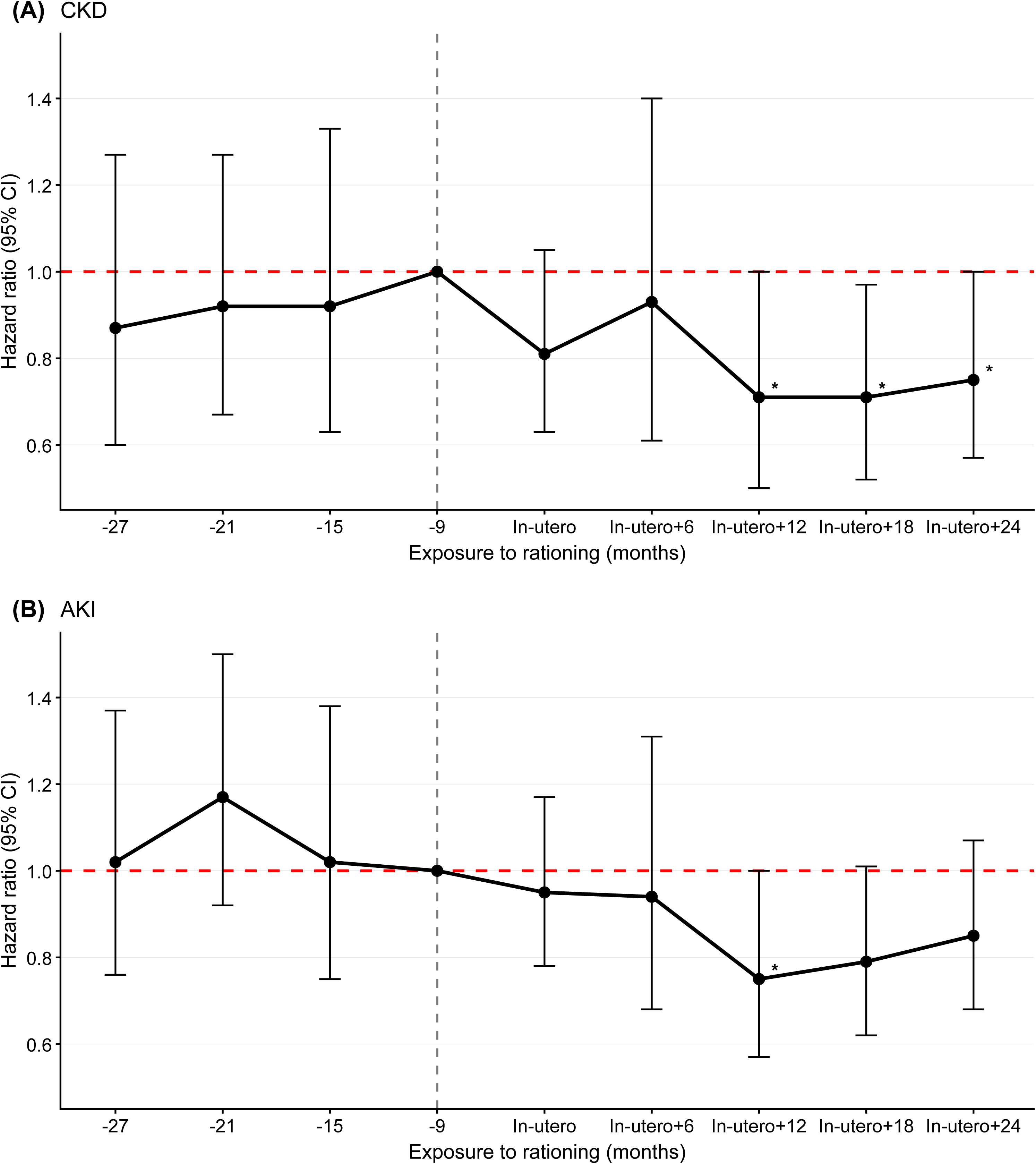
Hazard Ratios for Incident Kidney Diseases According to Duration of Early-Life Sugar Rationing Exposure ^#^. Hazard ratios (HRs) and 95% confidence intervals for **(A)** chronic kidney disease and **(B)** acute kidney injury based on Cox proportional hazards models are shown by birth date relative to the end of UK sugar rationing in September 1953. reference group was July to December 1954. The reference group was participants born July–December 1954. The vertical dashed line indicates the end of sugar rationing. Error bars represent 95% confidence intervals. * /+ indicates significance at p value smaller than 0.05/0.1. **^#^** Model adjusted for sex, ethnicity, birthplace (England versus Wales versus Scotland), deciles of north and east place-of-birth coordinates, calendar month of birth, real food prices (adjusted for Consumer Price Index), survey year, parental cardiovascular disease and diabetes, genetic risk score.

To enhance statistical power, all never-rationed participants were pooled into a single reference group. Compared with this group, those rationed in utero and through age 2 had a 22% lower risk of CKD (HR = 0.78, 95% CI: 0.66–0.93). For AKI, those rationed in utero and through the first postnatal year or through age 2 had a 21–22% lower risk (HR = 0.78–0.79). Rationing exposure was significantly associated with lower risks of both CKD (HR=0.84) and AKI (HR=0.82) (**Table 2**). Sensitivity analyses were consistent with primary findings (**Table S6**).

**Table 2.**
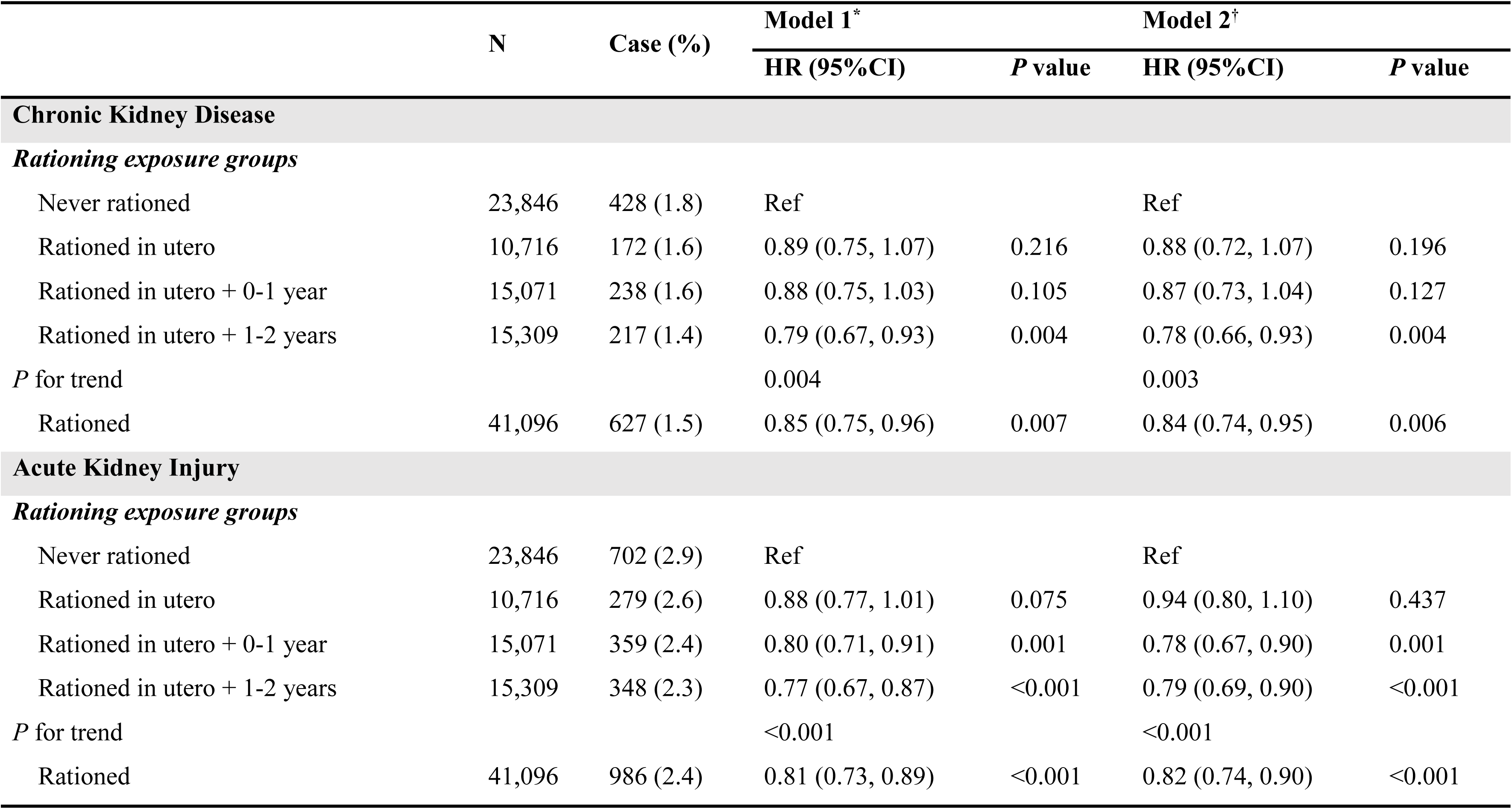
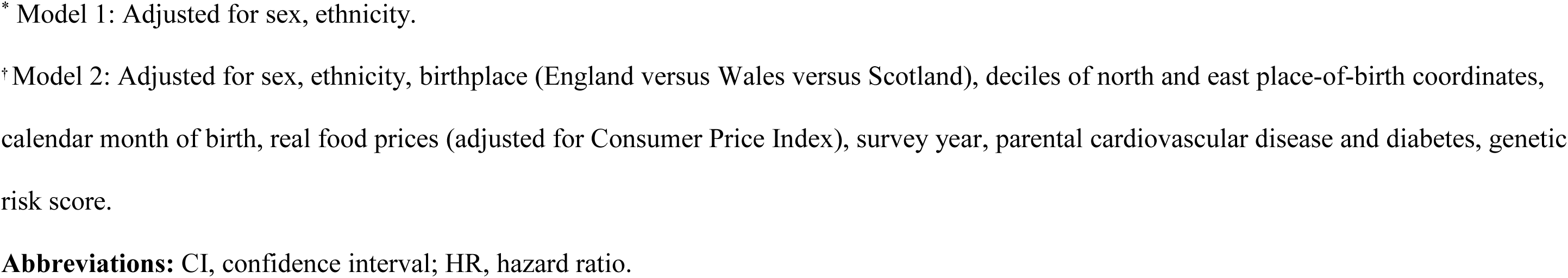
Associations of Early-Life Sugar Rationing Exposure with Incident Kidney Diseases.

|  | N | Case (%) | Model 1* |  | Model 2† |  |
| --- | --- | --- | --- | --- | --- | --- |
|  |  |  | HR (95%CI) | P value | HR (95%CI) | P value |
| Chronic Kidney Disease |  |  |  |  |  |  |
| Rationing exposure groups |  |  |  |  |  |  |
| Never rationed | 23,846 | 428 (1.8) | Ref |  | Ref |  |
| Rationed in utero | 10,716 | 172 (1.6) | 0.89 (0.75, 1.07) | 0.216 | 0.88 (0.72, 1.07) | 0.196 |
| Rationed in utero + 0-1 year | 15,071 | 238 (1.6) | 0.88 (0.75, 1.03) | 0.105 | 0.87 (0.73, 1.04) | 0.127 |
| Rationed in utero + 1-2 years | 15,309 | 217 (1.4) | 0.79 (0.67, 0.93) | 0.004 | 0.78 (0.66, 0.93) | 0.004 |
| P for trend |  |  | 0.004 |  | 0.003 |  |
| Rationed | 41,096 | 627 (1.5) | 0.85 (0.75, 0.96) | 0.007 | 0.84 (0.74, 0.95) | 0.006 |
| Acute Kidney Injury |  |  |  |  |  |  |
| Rationing exposure groups |  |  |  |  |  |  |
| Never rationed | 23,846 | 702 (2.9) | Ref |  | Ref |  |
| Rationed in utero | 10,716 | 279 (2.6) | 0.88 (0.77, 1.01) | 0.075 | 0.94 (0.80, 1.10) | 0.437 |
| Rationed in utero + 0-1 year | 15,071 | 359 (2.4) | 0.80 (0.71, 0.91) | 0.001 | 0.78 (0.67, 0.90) | 0.001 |
| Rationed in utero + 1-2 years | 15,309 | 348 (2.3) | 0.77 (0.67, 0.87) | <0.001 | 0.79 (0.69, 0.90) | <0.001 |
| P for trend |  |  | <0.001 |  | <0.001 |  |
| Rationed | 41,096 | 986 (2.4) | 0.81 (0.73, 0.89) | <0.001 | 0.82 (0.74, 0.90) | <0.001 |

\* Model 1: Adjusted for sex, ethnicity.
† Model 2: Adjusted for sex, ethnicity, birthplace (England versus Wales versus Scotland), deciles of north and east place-of-birth coordinates, calendar month of birth, real food prices (adjusted for Consumer Price Index), survey year, parental cardiovascular disease and diabetes, genetic risk score.
**Abbreviations:** CI, confidence interval; HR, hazard ratio.

### Stratified Analysis

The inverse associations between early-life sugar rationing and kidney disease risk were consistent across all subgroups examined, with no evidence of effect modification by sex, ethnicity, birthplace, parental history of diabetes or cardiovascular disease, or polygenic risk for kidney diseases (dichotomized at the median; all *P* for interaction > 0.05; **Figures S5–S6**).

### Contemporaneous Negative Control Analysis

Baseline characteristics of the negative control populations are shown in **Tables S7 and S8**. Among non-UK-born UK Biobank participants (n = 3,459), there was no association between birth timing relative to the rationing period and kidney outcomes (all *P* > 0.05; **Table S9**). Similarly, in the CHARLS cohort (n = 3,911), birth after the rationing period was not significantly associated with kidney outcomes (HR = 0.86, 95% CI: 0.72–1.02; **Table S10**).

### Mediation Analyses

#### Phenotype mediation

After excluding phenotypes with >20% missing data, 74 of 81 phenotypes were retained for analysis (**Table S11**). Multivariable linear regression with FDR correction identified 50 phenotypes associated with sugar rationing (**Table S12, Figure S7**). Subsequent Cox proportional hazards models identified phenotypes significantly associated with kidney disease risk, which were retained as candidate mediators (**Table S13**).

Basal metabolic rate, a cardio-metabolic indicator, mediated 8.95% of the CKD association and 9.24% of the AKI association. Mediation analyses also identified adiposity-related traits as significant mediators (fat-free mass proportion mediated: 8.90% for CKD, 9.18% for AKI). Other metabolic mediators included apolipoprotein A (5.01% for CKD, 5.05% for AKI), total cholesterol (4.61% for CKD, 4.70% for AKI), LDL cholesterol (3.15% for CKD, 3.23% for AKI), and apolipoprotein B (2.12% for CKD, 2.20% for AKI). Blood parameters (haemoglobin, haematocrit, red blood cell count) mediated 1-2.5% of the total effect, while the systemic immune-inflammation index showed minimal mediation (<1%) (**Figure 3, Table S14**).

**Figure 3.**
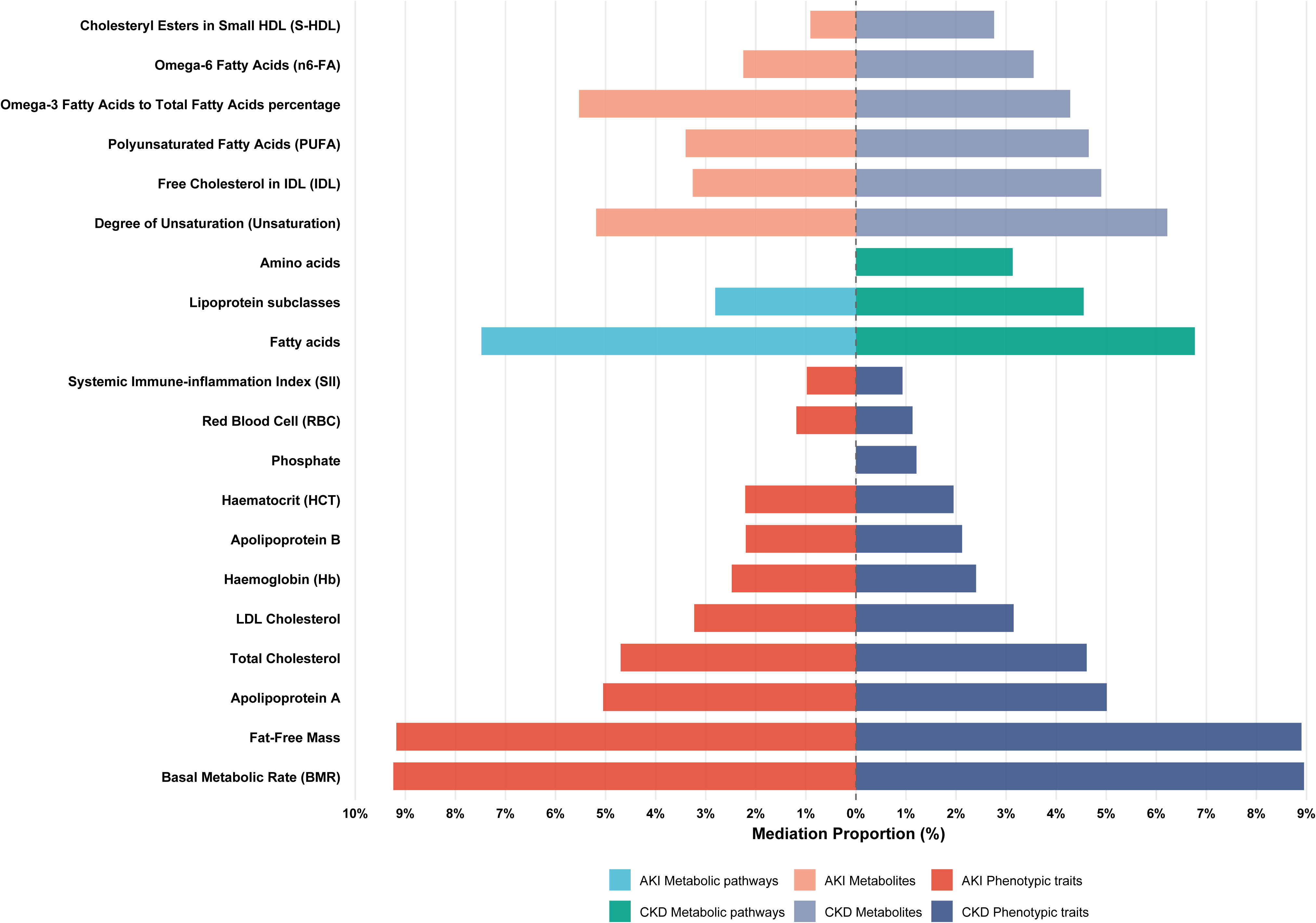
Mediated Proportion of the Association Between Early-Life Sugar Rationing and Incident Kidney Diseases by Phenotypic Traits, Metabolite Categories, and Individual Metabolites. Bars represent the percentage of the total protective association mediated by each factor (1,000 bootstrap iterations), separately for CKD and AKI. Mediators include phenotypic traits (e.g., BMR, fat-free mass, lipids), metabolite categories (fatty acids, lipoprotein subclasses, amino acids), and individual metabolites (e.g., PUFA, omega-3/omega-6, IDL free cholesterol). All models were adjusted for sex, ethnicity, birthplace (England vs. Wales vs. Scotland), deciles of north and east place-of-birth coordinates, calendar month of birth, real food prices (adjusted for Consumer Price Index), survey year, parental history of cardiovascular disease and diabetes, and genetic risk score for kidney outcomes.

#### Metabolomic mediation

LASSO regression identified 49 plasma metabolites significantly associated with early-life sugar rationing, representing eight major groups: lipoprotein subclasses, relative lipoprotein lipid concentrations, glycolysis-related metabolites, fatty acids, fluid balance, amino acids, lipoprotein particle sizes, and ketone bodies. Of these, 26 were positively associated and 23 inversely associated with rationing exposure (**Table S15**).

Mediation analyses evaluated whether metabolite categories and individual metabolites mediated the sugar rationing–kidney disease association.

Pathway-specific analysis identified three major mediating metabolic categories. Fatty acids mediated 6.77% and 7.48% of the total effect on CKD and AKI risk, respectively; lipoprotein subclasses mediated 4.55% (CKD) and 2.81% (AKI). Amino acids mediated 3.13% of the CKD association, but not significant of AKI association (**Table S14**).

Among the 49 metabolites, six exhibited significant mediating effects. For CKD, significant mediators included degree of unsaturation (6.22%), free cholesterol in intermediate-density lipoprotein (IDL) (4.90%), polyunsaturated fatty acids (PUFA) (4.65%), omega-3 fatty acids percentage (4.28%), omega-6 fatty acids (3.55%), and cholesteryl esters in small high-density lipoprotein (S-HDL) (2.76%). For AKI, the same metabolites showed mediated proportions of 5.19%, 3.26%, 3.40%, 5.53%, 2.25%, and 0.91%, respectively (**Figure 3, Table S14**).

## Discussion

In this quasi-experimental study, sugar restriction during the first 1000 days of life was associated with significantly lower risks of CKD and AKI in adulthood in a clear dose–response manner. These associations were robust to sensitivity and negative control analyses and were partially mediated by favorable metabolic and lipid profiles, with a distinct metabolomic signature identified.

### Dose–response relationship and developmental programming

The graded association between duration of early-life sugar restriction and adult kidney disease risk supports a developmental programming effect [5,7]. The strongest protection was observed among individuals exposed across the full 1000-day window, consistent with the sensitivity of nephrogenesis and early metabolic programming to nutritional cues during this critical period [6]. Our findings extend prior work using the same natural experiment, which reported protection against diabetes and hypertension [16], by directly linking early-life sugar restriction to reduced risks of CKD and AKI—major long-term complications of metabolic disease. This dose–response pattern further strengthens causal inference and underscores that the timing and duration of early-life sugar exposure are critical determinants of lifelong kidney health.

### Metabolic signature of early-life sugar restriction

Large-scale metabolomic profiling revealed a distinct metabolic signature associated with early-life sugar restriction, characterized by a predominance of favorable lipid-related metabolites. Individuals exposed to rationing had higher levels of polyunsaturated fatty acids, omega-3 and omega-6 fatty acids, and higher degrees of fatty acid unsaturation, alongside lower levels of VLDL subfractions and triglycerides. This pattern—consistent with a metabolically efficient phenotype—suggests that early-life nutritional programming induces persistent shifts in lipid metabolism that extend into adulthood [37, 38]. These findings align with experimental evidence indicating that maternal high-sugar intake disrupts lipid homeostasis and promotes renal lipotoxicity in offspring [10, 39, 40].

### Mediating pathways

Mediation analyses identified metabolic efficiency and body composition as major mediators, with basal metabolic rate and fat-free mass accounting for approximately 9% of the protective effects. These findings support the hypothesis that early-life sugar restriction promotes lean mass development and metabolic efficiency, thereby reducing long-term kidney risk [41, 42]. Lipid metabolism constituted another key mediating domain, with fatty acid composition and lipoprotein subfractions explaining 4–7% of the associations. Notably, systemic inflammation showed negligible mediation, suggesting that the renoprotective effects operate primarily through metabolic reprogramming rather than anti-inflammatory pathways. The convergence of evidence from clinical phenotypes, metabolic pathways, and individual metabolites provides a coherent mechanistic framework linking early-life nutrition to lifelong kidney health [43].

### Negative control analyses

The absence of associations in non-UK-born UK Biobank participants and the CHARLS cohort—populations not exposed to the UK rationing policy—argues against residual confounding by secular trends or unmeasured factors [16,20]. These negative control analyses further support the causal interpretation of the main findings.

### Clinical and public health implications

Our findings have direct implications for clinical practice and public health. Sugar intake during rationing approximated current WHO recommendations, supporting the relevance of these findings to contemporary settings. Clinically, limiting added sugar during pregnancy and early childhood may confer lasting kidney protection; children with high early-life sugar exposure may warrant targeted monitoring of UACR and eGFR, particularly when facing AKI precipitants such as major surgery or nephrotoxic drugs. For public health, these findings support policies to reduce added sugar consumption, including front-of-package warning labels and restrictions on child-targeted marketing.

### Limitations

Several limitations warrant consideration. First, exposure was defined by birthdate, not individual sugar intake, potentially causing nondifferential misclassification. Second, despite extensive adjustment, residual confounding from unmeasured secular changes (e.g., post-war economic recovery) or other in-utero factors (e.g., maternal stress, overall diet) remain possible. Third, metabolomic measurements were obtained only in adulthood, precluding life-course assessment. Fourth, survivor bias may affect our estimates if early high sugar exposure increased childhood mortality, though competing-risk analyses gave consistent results. Fifth, we assumed no migration during the first 1000 days; any such misclassification is likely minimal in this birth cohort. Sixth, generalizability to other ancestries and contemporary settings requires further study. Finally, although negative control analyses argue against secular confounding, they cannot rule out all unmeasured confounding specific to the UK-born cohort.

## Conclusions

In this natural experiment, early-life sugar restriction during the first 1000 days was associated with dose-dependent reductions in CKD and AKI risk in adulthood, partially mediated by favorable body composition, metabolic efficiency, and lipid profiles. These findings identify early-life sugar exposure as a modifiable programming factor for lifelong kidney health and support public health strategies to reduce added sugar during pregnancy and early childhood. Future research should explore epigenetic mechanisms and test sugar-reduction interventions in randomized trials.

## Declarations

### Contributors

Xianglian Cai, Yuanyuan Zhang, and Xianhui Qin conceived and designed the study. Xianglian Cai, Xiaolong Liang, Dan Chen, Yiwei Zhang, and Ziliang Ye performed the data management, statistical analyses, and had full access to the underlying data. Xianglian Cai and Xianhui Qin accessed and verified the data. Xianhui Qin acquired funding and supervised the project. Xianglian Cai, Yuanyuan Zhang, and Xianhui Qin drafted the manuscript. All authors reviewed and edited the manuscript for important intellectual content and approved the final version. All authors had full access to all the data in the study and had final responsibility for the decision to submit for publication.

### Data sharing

De-identified participant data from UK Biobank and CHARLS are available upon bona fide application. UK Biobank: https://www.ukbiobank.ac.uk/ (proposal approval and data access agreement required). CHARLS: http://charls.pku.edu.cn/ (registration required). A data dictionary is provided within each platform. The statistical analysis plan and analytic code are available from the corresponding author upon request, beginning at publication, for replication purposes. Access to code requires a signed data access agreement.

### Ethical approval

The UK Biobank received ethical approval from the North West Research Ethics Committee (11/NW/0382). The CHARLS was approved by the Ethical Review Committee of Peking University (IRB00001052-11015) and the Biomedical Ethics Review Committee of Peking University. All participants provided written informed consent.

## Data Availability

De-identified participant data from UK Biobank and CHARLS are available upon application and approval. UK Biobank: https://www.ukbiobank.ac.uk/ (proposal approval and data access agreement required). CHARLS: http://charls.pku.edu.cn/ (registration required). A data dictionary is provided within each platform. The statistical analysis plan and analytic code are available from the corresponding author upon reasonable request for replication purposes.

## Acknowledgments

This research was funded by the National Natural Science Foundation of China (grants 82570914, 81973133, 82030022, 82330020), Key Technologies R&D Program of Guangdong Province (2023B1111030004), Guangdong Provincial Clinical Research Center for Kidney Disease (2020B1111170013), the Program of Introducing Talents of Discipline to Universities, 111 Plan (D18005), and President Foundation of Nanfang Hospital, Southern Medical University (2024B029). The funders had no role in study design, data collection, analysis, interpretation, or manuscript writing.

We thank all participants of the UK Biobank and CHARLS, as well as their respective staff and investigators for their invaluable contributions. This research was conducted using the UK Biobank Resource under Application Number 73201.

No medical writer or editor was involved in the creation of this manuscript. AI-assisted technologies were used solely for language improvement. The authors assume full responsibility for the content.

